# Corticosteroids for improving patient relevant outcomes in HELLP syndrome: a systematic review and meta-analysis

**DOI:** 10.1101/2023.10.28.23297641

**Authors:** Asmaa F Kasem, Hamdy B Alqenawy, Marwa A Elgendi, Radwa R Ali, Rania HM Ahmed, Mohammad N Sorour, Khadiga MH Hegab, Rania G El-skaan, Rowyna H El Helw, Mohamed S Elsewefy, Maya M Abdelrazek, Yasser M Elrefaey, Mohamed YG Albahaie, Mohamed H Salama, Ashraf F Nabhan

## Abstract

**Objectives:** We conducted an update of systematic review to assess the effects of corticosteroids vs placebo or no treatment for improving patient relevant outcomes in hemolysis, elevated liver enzymes and low platelets (HELLP) syndrome.

**Design:** A systematic review and meta-analysis of randomized controlled trials.

**Data sources:** CENTRAL, MEDLINE/PubMed, Web of Science, and Scopus from the date of inception of the databases to September 20, 2023. The reference lists of included studies and other systematic reviews were thoroughly searched.

**Eligibility criteria:** We included randomized controlled trials that enrolled women with HELLP syndrome, whether antepartum or postpartum, to receive any corticosteroid versus placebo or no treatment. No restrictions on language or date of publication were made.

**Data extraction and synthesis:** We used a dual independent approach for screening titles and abstracts, full text screening, data extraction and risk of bias assessment. Pairwise meta-analyses were conducted, where two or more studies met methodological criteria for inclusion. The Grading of Recommendations, Assessment, Development and Evaluation approach was used to assess certainty for the pre-specified important outcomes.

**Results:** Fifteen trials (821 women) compared corticosteroids with placebo or no treatment. The effect of corticosteroids is uncertain for the primary outcome i.e., maternal death (risk ratio 0.77, 95% confidence interval 0.25 to 2.38, very low certainty evidence). The effect of corticosteroids is also uncertain for other important outcomes including pulmonary edema, dialysis, liver morbidity (hematoma, rupture, and failure), or perinatal death because of very low certainty evidence. Low certainty evidence suggests that corticosteroids have little or no effect on the need for platelet transfusion (risk ratio 0.9821; 95% confidence interval 0.6031 to 1.5994) and may result in a slight reduction in acute renal failure (risk ratio 0.6658; 95% confidence interval 0.3965 to 1.1179)

**Conclusion:** In women with HELLP syndrome, the effect of corticosteroids vs placebo or no treatment is uncertain for patient relevant outcomes including maternal death, maternal morbidity, and perinatal death.

**Article summary:** *Strengths and limitations of this study:*

- We used robust systematic review and meta-analysis methods.
- We synthesized results on patient relevant outcomes that are critical for decision making.
- The available evidence was associated with limitations related to the small sample size of included trials, different timing to initiate corticosteroids administration, and reporting bias.
- Outcomes assessed using the Grading of Recommendations, Assessment, Development and Evaluation framework were judged to have very low certainty of evidence due to extremely serious imprecision.

## Introduction

The syndrome of hemolysis, elevated liver enzymes and low platelets (HELLP) has an incidence of 2.5 per 1000 singleton deliveries and it complicates 20% of women diagnosed with severe pre-eclampsia. [1,2]

The pathophysiology of HELLP syndrome that is usually diagnosed between 27 and 37 weeks, is not completely understood. [3]

The diagnosis depends on laboratory findings of microangiopathic hemolysis, thrombocytopenia, and elevated liver enzymes. Different investigators reported different threshold of hematologic and biochemical values for diagnosis of the syndrome or for determining the prognosis. [4,5]

The presence of HELLP syndrome is associated with significant maternal mortality and morbidity including acute renal and liver failure. [1] Approximately 70% of pregnancies complicated by HELLP syndrome require preterm delivery, thus increasing perinatal morbidity and mortality. [5]

Observational studies suggested that steroid treatment in HELLP syndrome may improve disordered maternal hematological and biochemical features and perhaps perinatal mortality and morbidity. Clinical trials examined the effects of corticosteroids for the treatment of maternal HELLP syndrome. Various regimens have been reported using prednisolone, dexamethasone, or betamethasone. [2,5–7]

Current practice and clinical guidelines require an updated evidence synthesis because the latest available synthesis was published in 2010, [8] new studies have been published, and the clinical question remains relevant to decision makers.

We conducted this systematic review to update the synthesized evidence regrading the effects of corticosteroids versus placebo or no treatment for improving outcomes in women with HELLP syndrome.

## Methods

### Protocol and registration

This systematic review was conducted following the methodological standards of Cochrane Handbook. [9] We prospectively registered the protocol in Open Science Platform. The full text of the protocol is available in an open access registry and as an online as Supplemental file 1. We reported the review using the Preferred Reporting Items for Systematic reviews and Meta-Analyses (PRISMA) standards. [10] The full checklist is available as Supplemental file 2.

### Eligibility criteria

We included published randomized controlled trials that recruited women with HELLP syndrome, confirmed by objective testing. We included studies comparing corticosteroids versus placebo or no treatment. The primary outcome measure was maternal death. Other outcomes included accute pulmonary edema; acute renal failure; dialysis, liver morbidity (hematoma, ruptured liver, and failure), need for platelet transfusion, and perinatal death.

### Information sources

A comprehensive literature search was initially conducted on September 20, 2023. We did not impose language or other restrictions on any of the searches. We searched bibliographic databases (Cochrane Central Register of Controlled Trials (CENTRAL), MEDLINE/PubMed) and citation indexes (Web of Science and Scopus). We included the terms (HELLP Syndrome) AND (corticosteroids or glucocorticoids or Dexamethasone or Betamethasone or Prednisolone). The detailed exact strategy adapted for each database is provided in Supplemental file 3 and is available as an open access registry document. We peer-reviewed the search strategy and further tested it with a set of known relevant, ‘gold standard’, reports. We also searched clinical trial registries (ClinicalTrials.gov and the World Health Organization International Clinical Trials Registry Platform) to identify ongoing trials. We finally searched reference lists and explored the cited-by logs of identified studies and previously published reviews.

### Study selection

All reports identified in the databases were imported to Bibtex library using Jabref version 5. After removing duplicates, two authors independently screened all titles and abstracts for eligibility. We retrieved and assessed the full text of all reports that potentially met our eligibility criteria during screening. Two authors (AFK, HBA, MAE, RHA) independently assessed each full-text article. Disagreements regarding trial eligibility was resolved by consensus and finally resolved by a third author (AFN).

### Data collection process

For eligible studies, we extracted the data in duplicates using an offline electronic form. We resolved discrepancies through discussion. Extracted data were transcribed to a spreadsheet and checked for accuracy. We contacted authors of the original reports, if needed, to provide details regarding unclear or missing data.

### Data items

Extracted data included study design, sample size, description of included participants, description of the intervention, outcomes, trial registration, and funding sources, and country.

### Study risk of bias assessment

Two authors (AFK, HBA, MAE, RHA) independently used the Risk of Bias 2 (RoB 2) tool to assess the risk of bias of study results contributing information to each of the outcomes specified for inclusion in the Summary of Findings table.

We assessed the following risk of bias domains as outlined in Cochrane Handbook for Systematic Reviews of Interventions: 1) risk of bias arising from the randomization process; 2) risk of bias due to deviations from the intended interventions (effect of assignment to intervention); 3) risk of bias due to missing outcome data; 4) risk of bias in measurement of the outcome; and 5) risk of bias in selection of the reported result. Each domain was judged as being at “low risk of bias”, “some concerns”, or “high risk of bias”. Trials with “low risk of bias” in all domains were classified as being at overall “low risk of bias”. RCTs with one domain judged to be at “some concerns”, but no domain judged to be at “high risk of bias”, were classified as being at overall “some concerns” of risk of bias. RCTs were classified as being at overall “high risk of bias” if at least one domain was judged as being at “high risk of bias”. However, if a trial was judged to be at “some concerns” due to risk of bias for multiple domains, it was judged as being at overall “high risk of bias” if the assessors judged that the multiple concerns amounted to a serious risk of bias. In case of discrepancies among their judgments and inability to reach consensus, we consulted the senior author (AFN) to reach a final decision.

### Effect measures

For dichotomous data, we presented results as summary risk ratio (RR) with 95% confidence intervals (CI). None of the outcomes of interest were meta-analysed as a continuous variable. The unit of analysis was the individual participant. We used a complete case approach for analysis. Data related to participants reported as not compliant was analyzed on an intention-to-treat basis.

### Synthesis methods

Fixed-effect meta-analysis was performed to combine data of trials that are judged to be sufficiently similar in terms of intervention, populations, and methods. We planned to investigate substantial statistical heterogeneity, defined as I^2^ statistic ≥ 50% or P <0.1.

We performed the planned subgroup analysis by gestational age at enrollment (ante-vs postpartum) and by type of corticosteroids. We assessed subgroup differences by interaction tests. Results of the subgroup analyses were reported by mentioning the Chi^2^ statistic and P value, and the interaction test I^2^ value.

Sensitivity analysis was performed to explore robustness of pooled estimate using outcome data from trials with a low risk of bias.

Synthesis was performed using RStudio 2023.06.1 Build 524 (MacOS, Apple Silicon version), R 4.3.1 (2023-06-16) [11] and R package meta version 6.5. [12]

### Reporting bias assessment

We explored whether the study was included in a trial registry and whether a protocol was available. We planned to examine funnel plots to assess the potential for publication bias if we found 10 or more studies reporting on a particular outcome.

### Certainty assessment

We used the Grading of Recommendations, Assessment, Development and Evaluation (GRADE) approach to create the Summary of Findings table. [13] Briefly, GRADE uses study limitations, consistency of effect, imprecision, indirectness, and publication bias to assess the certainty of evidence for each outcome. A summary of the intervention effect and a measure of certainty was produced using the GRADE Profiler Guideline Development Tool (GRADEpro GDT) software [14] for the prespecified important outcomes: maternal death, pulmonary edema, renal failure, dialysis, liver morbidity, need for platelet transfusion, and perinatal death. One author (A.N.) conducted GRADE assessments and the decisions on downgrading. This was discussed for final approval by all authors.

### Patient and public involvement

We consulted with patients by asking a group to review and comment on an early draft of the manuscript.

## Results

### Study selection

Bibliographic database search identified 154 records. After removing duplicates, 86 titles and abstracts were screened. Twenty four titles required further assessment. One is an ongoing CTRI/2020/12/029730 and the full-text reports of twenty three published reports were assessed using the predefined eligibility criteria. We excluded two reports identified in our search. One study did not meet our inclusion criteria for participants as it enrolled women with low platelets not HELLP syndrome. The other study was terminated because of the inability to recruit the required sample. Fifteen studies (21 reports published between 1994 and 2019) including 821 women were found eligible Figure 1.

**Figure 1:**
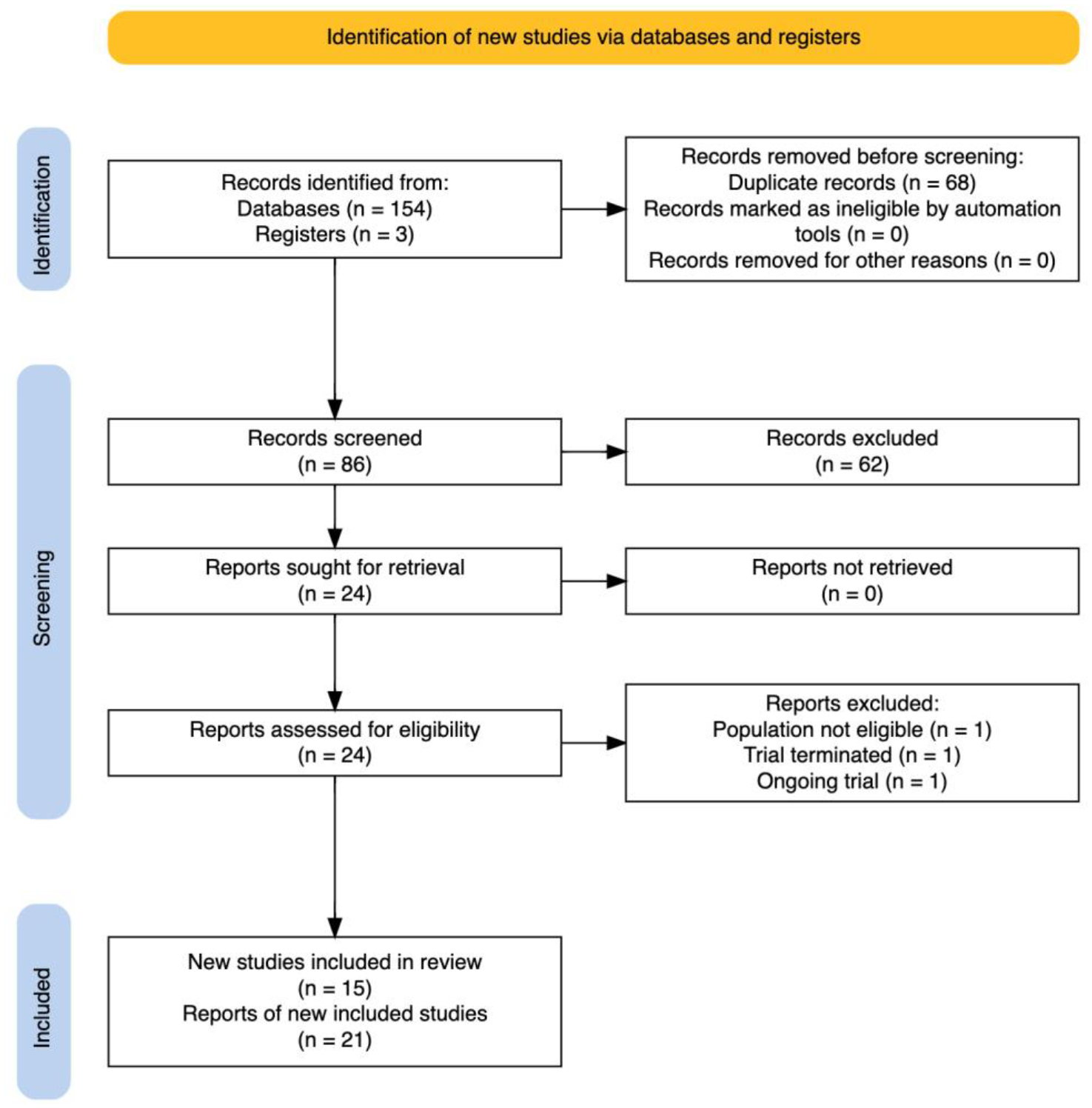
PRISMA flow diagram. PRISMA, Preferred Reporting Items for Systematic reviews and Meta-Analyses

### Study characteristics

We summarized the characteristics of the included studies in Table 1.

**Table 1:**
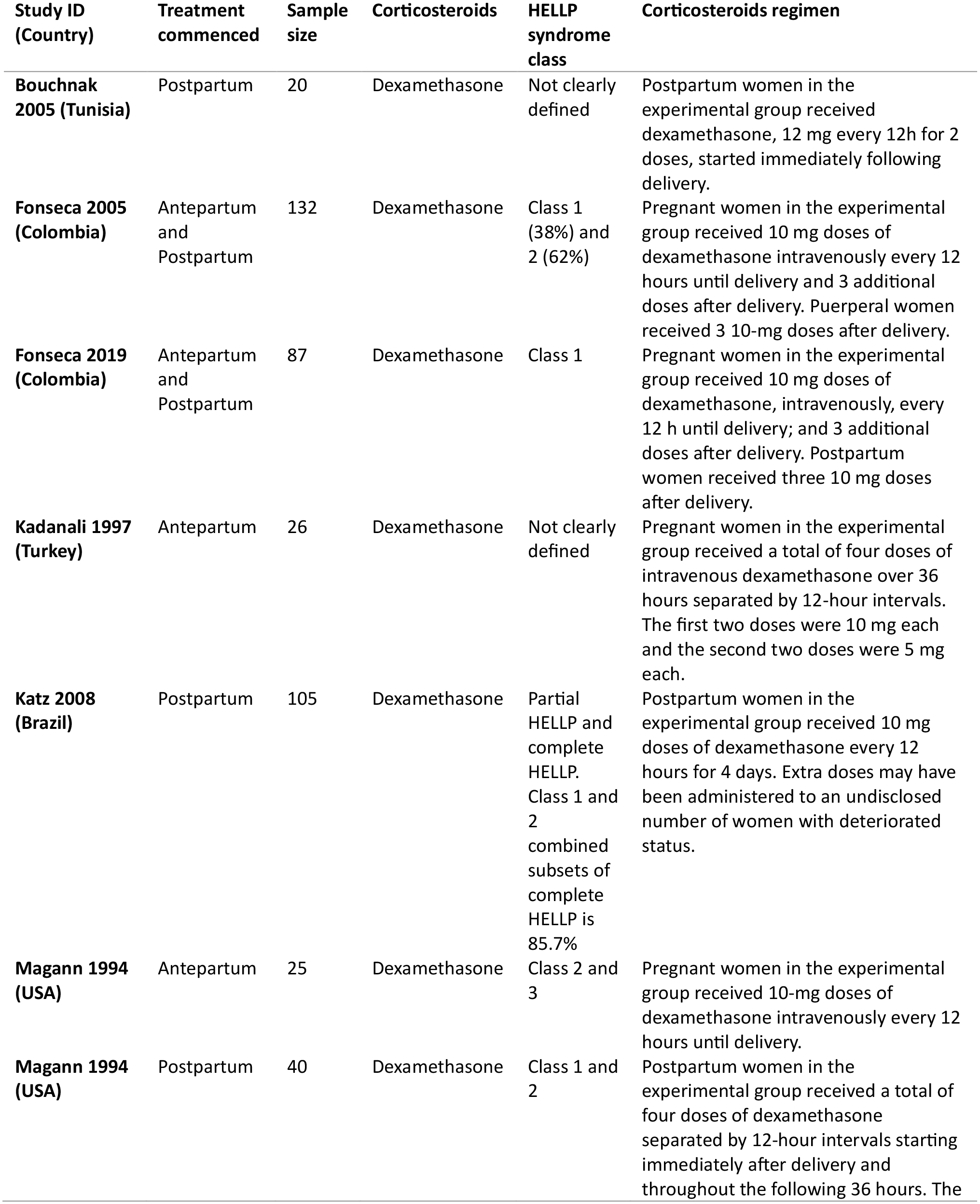
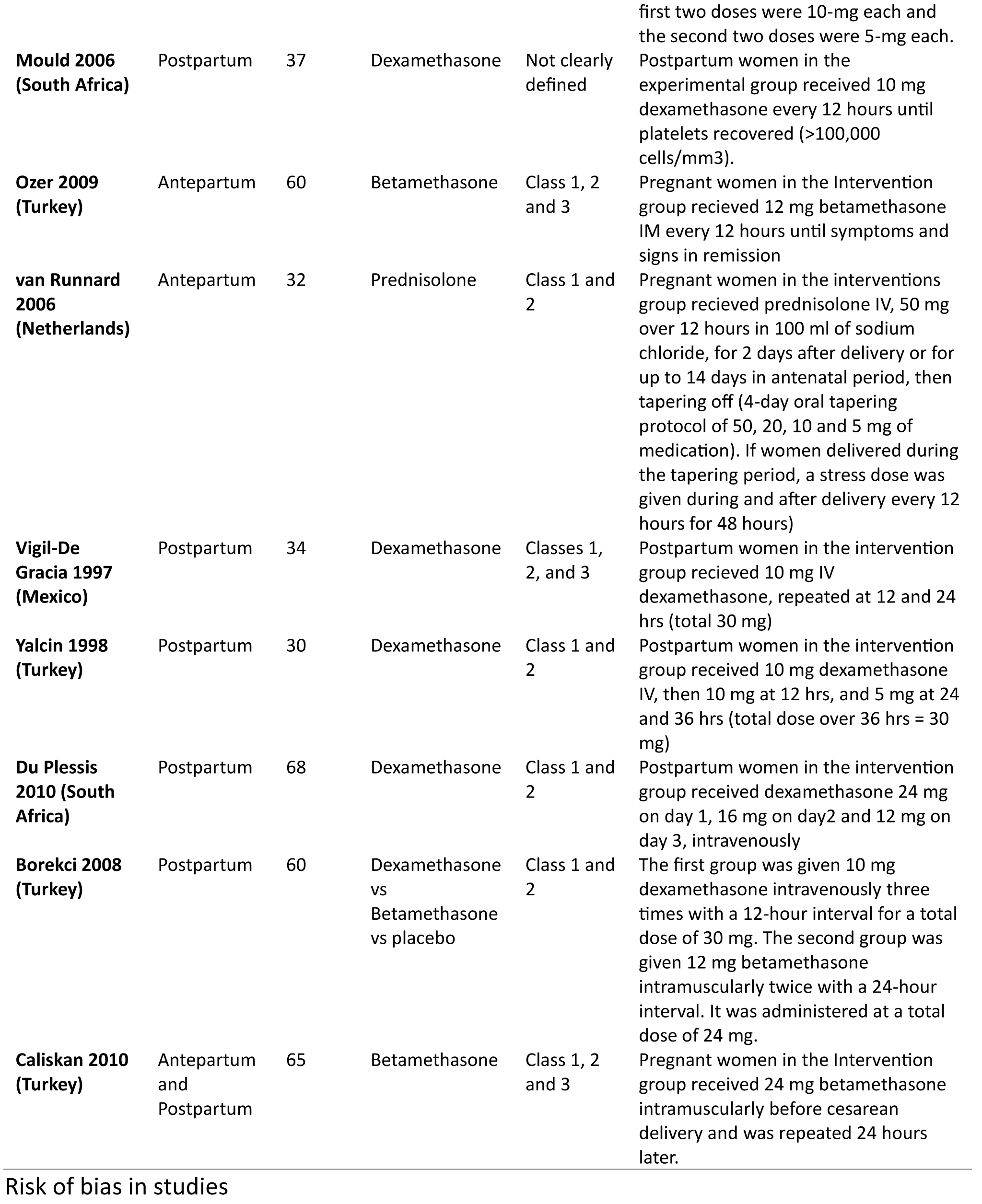
Characteristics of included studies.

The fifteen included trials recruited 821 women with HELLP syndrome. Criteria for recruitment in five trials [15–20] were a diagnosis of HELLP class 1 or 2 on the Mississippi HELLP classification system. One trial exclusively recruited women with class 1. [21] One trial recruited women with class 2 and 3. [22] Three studies [23–25] recruited women with HELLP classes 1, 2, and 3. One study [26] included women with partial HELLP (1 or more parameters abnormal) (61/105 [58.1%]) and complete HELLP (all parameters abnormal) (44/105 [41.9%]); class 1 and 2 combined subset accounted for 85.7% of participants with complete HELLP. Three studies [27,28,29] did not report explicitly on the class of HELLP syndrome.

Eleven trials administered dexamethasone vs placebo or no treatment, [16–18,20–22,25–29] two trials administered betamethasone, [23,24] and one trial administered prednisolone. [19] One multiple-arms trial compared dexamethasone vs betamethasone vs no treatment. [15]

Corticosteroids administration commenced after delivery in eight trials, [15,16,18,20,25–27,29] before delivery in five trials, [19,22–24,28] and in two trials [17,21] treatment commenced according to timing of recruitment whether before or after delivery.

### Risk of bias in studies

We assessed the risk of bias for the included RCTs contributing results to our outcomes using the RoB 2 tool. The overall risk of bias for all study results per outcome are available in Supplemental file 4.

The results of risk of bias assessments for the primary outcome, maternal death are depicted in Supplemental file 4. Overall, 5 out of 6 studies were judged overall to be at “low risk” of bias. One study [25] was judged to be at “high risk” of bias. The main reasons for having “high risk” in domain 1 were lack of description in the randomization process with baseline differences in platelet count between intervention groups that suggest a problem with the randomization process. There were “some concerns” in two other domains. First, people delivering the interventions were probably aware of participants’ assigned intervention during the trial. Second, there were no information whether the data that produced this result were analysed in accordance with a pre-specified analysis plan that was finalized before unblinded outcome data were available for analysis.

### Syntheses of results

#### Maternal death

Six trials (449 women) reported maternal death. The risk ratio (RR) was 0.77 (95% confidence intervals (CI) 0.25 to 2.38, Figure 2). The effect of any corticosteroid vs placebo or no treatment is uncertain. We downgraded the certainty of the evidence to very low due to extremely serious imprecision, Table 2.

**Figure 2:**
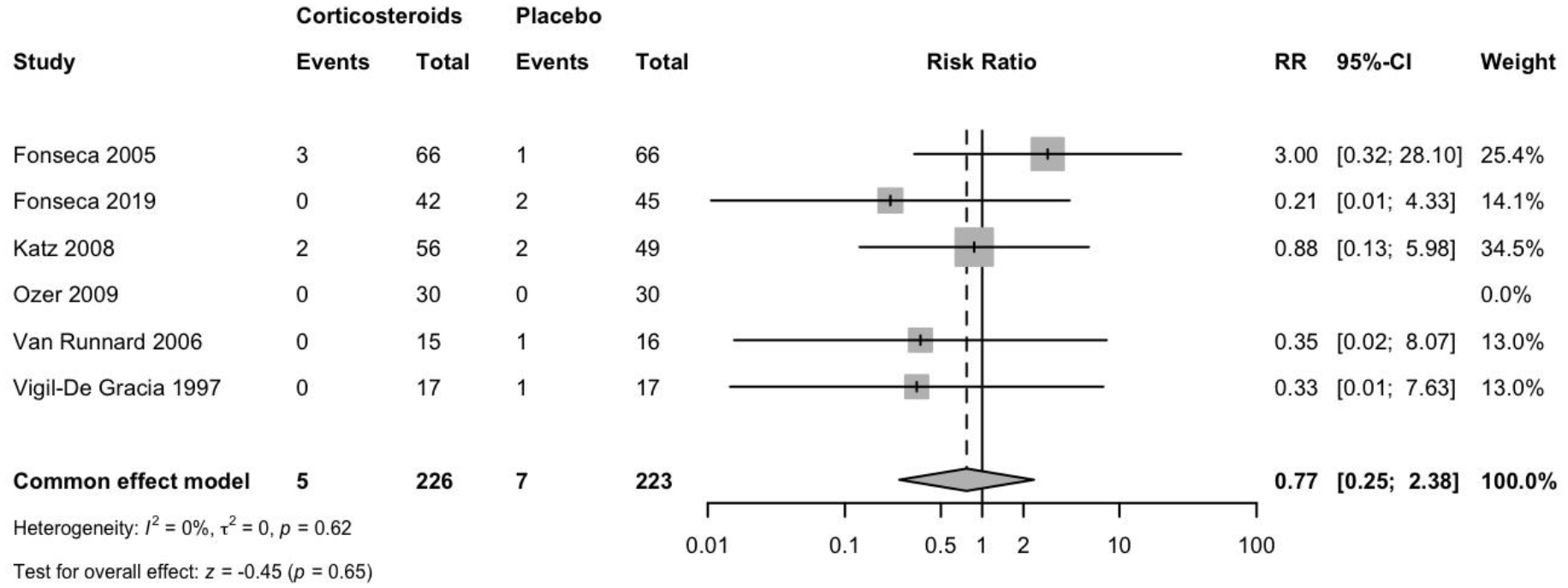
Maternal death: Corticosteroids vs placebo or no treatment

**Table 2:**
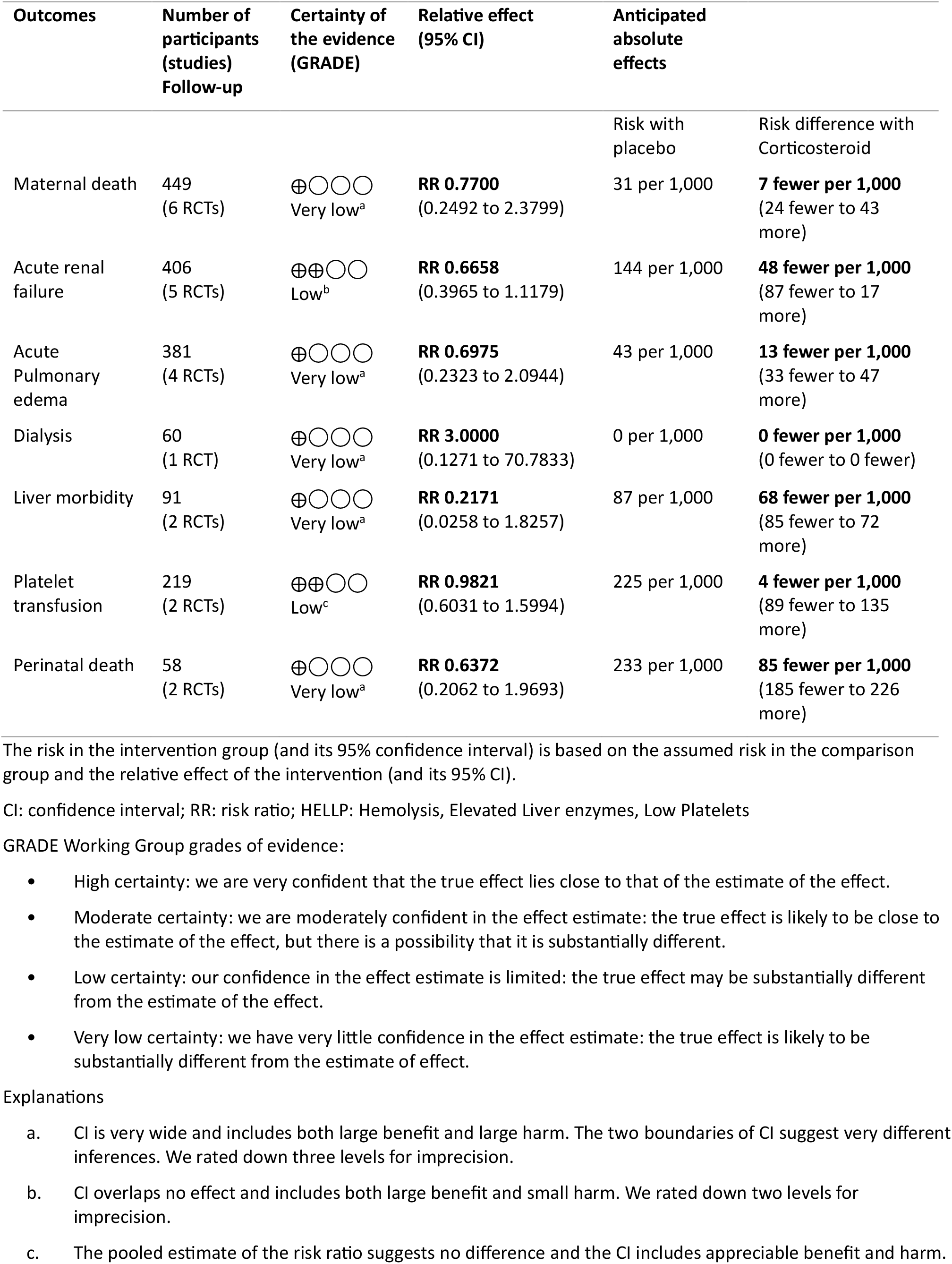
Summary of Findings: Corticosteroids compared to placebo for women with HELLP syndrome.

| Outcomes | Number of participants (studies) Follow-up | Certainty of the evidence (GRADE) | Relative effect (95% CI) | Anticipated absolute effects |  |
| --- | --- | --- | --- | --- | --- |
|  |  |  |  | Risk with placebo | Risk difference with Corticosteroid |
| Maternal death | 449 (6 RCTs) | ⊕○○○<br>Very low<sup>a</sup> | **RR 0.7700**<br>(0.2492 to 2.3799) | 31 per 1,000 | **7 fewer per 1,000**<br>(24 fewer to 43 more) |
| Acute renal failure | 406 (5 RCTs) | ⊕⊕○○<br>Low<sup>b</sup> | **RR 0.6658**<br>(0.3965 to 1.1179) | 144 per 1,000 | **48 fewer per 1,000**<br>(87 fewer to 17 more) |
| Acute Pulmonary edema | 381 (4 RCTs) | ⊕○○○<br>Very low<sup>a</sup> | **RR 0.6975**<br>(0.2323 to 2.0944) | 43 per 1,000 | **13 fewer per 1,000**<br>(33 fewer to 47 more) |
| Dialysis | 60 (1 RCT) | ⊕○○○<br>Very low<sup>a</sup> | **RR 3.0000**<br>(0.1271 to 70.7833) | 0 per 1,000 | **0 fewer per 1,000**<br>(0 fewer to 0 fewer) |
| Liver morbidity | 91 (2 RCTs) | ⊕○○○<br>Very low<sup>a</sup> | **RR 0.2171**<br>(0.0258 to 1.8257) | 87 per 1,000 | **68 fewer per 1,000**<br>(85 fewer to 72 more) |
| Platelet transfusion | 219 (2 RCTs) | ⊕⊕○○<br>Low<sup>c</sup> | **RR 0.9821**<br>(0.6031 to 1.5994) | 225 per 1,000 | **4 fewer per 1,000**<br>(89 fewer to 135 more) |
| Perinatal death | 58 (2 RCTs) | ⊕○○○<br>Very low<sup>a</sup> | **RR 0.6372**<br>(0.2062 to 1.9693) | 233 per 1,000 | **85 fewer per 1,000**<br>(185 fewer to 226 more) |

The risk in the intervention group (and its 95% confidence interval) is based on the assumed risk in the comparison group and the relative effect of the intervention (and its 95% CI).
CI: confidence interval; RR: risk ratio; HELLP: Hemolysis, Elevated Liver enzymes, Low Platelets
GRADE Working Group grades of evidence:
- High certainty: we are very confident that the true effect lies close to that of the estimate of the effect. - Moderate certainty: we are moderately confident in the effect estimate: the true effect is likely to be close to the estimate of the effect, but there is a possibility that it is substantially different. - Low certainty: our confidence in the effect estimate is limited: the true effect may be substantially different from the estimate of the effect. - Very low certainty: we have very little confidence in the effect estimate: the true effect is likely to be substantially different from the estimate of effect.
Explanations
- CI is very wide and includes both large benefit and large harm. The two boundaries of CI suggest very different inferences. We rated down three levels for imprecision. - CI overlaps no effect and includes both large benefit and small harm. We rated down two levels for imprecision. - The pooled estimate of the risk ratio suggests no difference and the CI includes appreciable benefit and harm.

The subgroup analysis did not show significant differences among groups whether by the timing of corticosteroid administration (test for subgroup differences P = 0.79) or by the type of corticosteroid (test for subgroup differences P = 0.60) (Supplemental file 4).

Sensitivity analysis to explore robustness of pooled estimate for maternal death, using outcome data from trials with a low risk of bias showed results similar to primary analysis with a RR 0.87 (95% CI 0.26 to 2.92) or by including studies with zero events with RR 0.79 (95% CI 0.27 to 2.32) (Supplemental file 4).

#### Acute pulmonary edema

The effect of any corticosteroid vs placebo or no treatment is very uncertain. Four trials (381 women) reported pulmonary edema. The risk ratio was 0.70 (95% confidence interval 0.23 to 2.09), Figure 3. We downgraded the certainty of the evidence to very low due to extremely serious imprecision.

**Figure 3:**
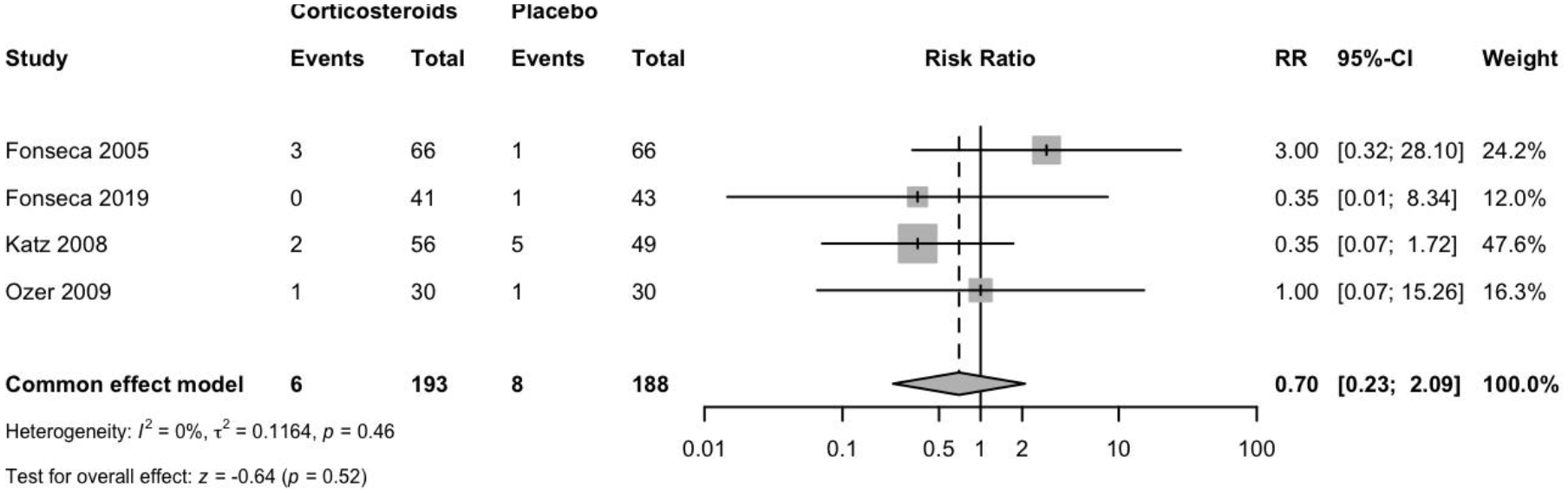
Acute pulmonary edema: Corticosteroids vs placebo or no treatment

#### Acute renal failure

Five trials (406 women) reported acute renal failure. Corticosteroids may result in a slight reduction in acute renal failure. The risk ratio was 0.6658 (95% CI 0.3965 to 1.1179), Figure 4. The certainty of evidence was low due to very serious imprecision.

**Figure 4:**
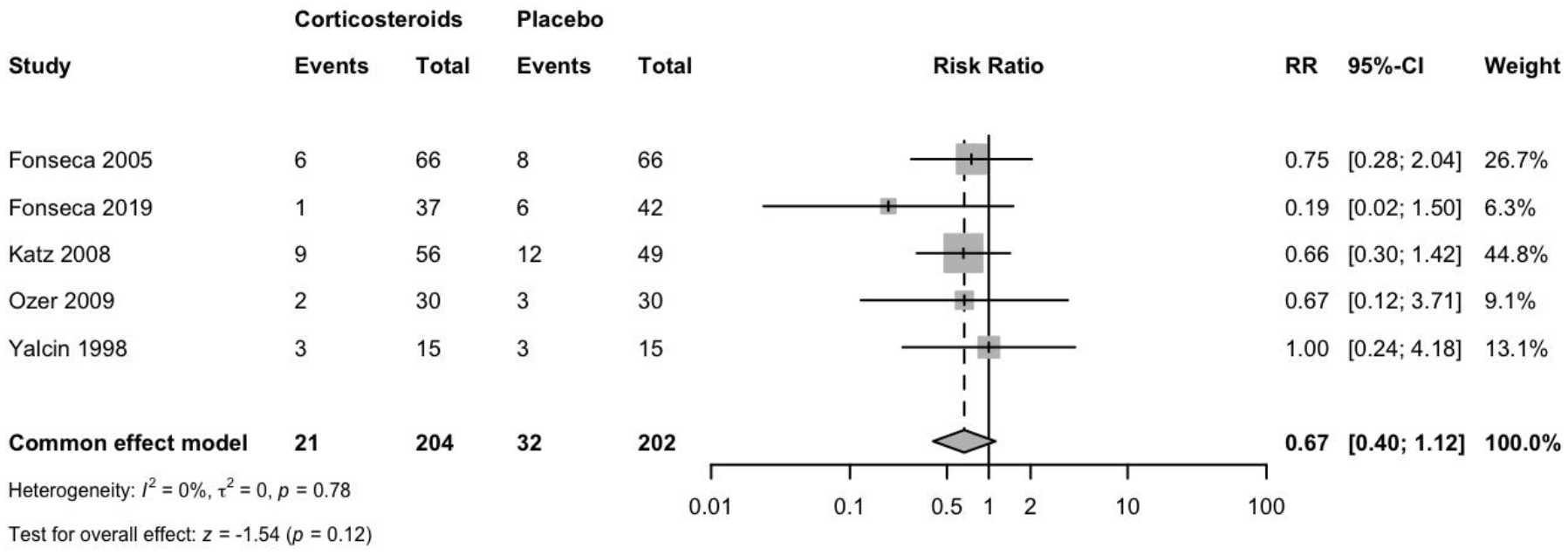
Acute renal failure: Corticosteroids vs placebo or no treatment

#### Dialysis

The evidence is very uncertain about the effect of corticosteroids on dialysis. The need for dialysis was reported in one study (60 women). The risk ratio was 3 (95% CI 0.1271 to 70.7833). The certainty of evidence was downgraded to very low due to extremely serious imprecision.

#### Liver morbidity (hematoma, rupture, or failure)

The evidence is very uncertain about the effect of corticosteroids on liver morbidity vs placebo or no treatment. Based on data from two studies (91 women), the risk ratio was 0.2171 (95% CI 0.0258 to 1.8257). We downgraded the certainty of evidence to very low due to extremely serious imprecision.

#### Platelet transfusion

Based on data from 219 women in two studies, corticosteroids have little or no difference in the need for platelet transfusion (RR 0.9821; 95% CI 0.6031 to 1.5994). We downgraded the certainty of evidence to low due to very serious imprecision.

#### Perinatal death

The evidence is very uncertain about the effect of corticosteroids on perinatal death. Based on data from two studies (58 women), the risk ratio was 0.6372 (95% CI 0.2062 to 1.9693). The certainty of evidence was very low due to extremely serious imprecision.

### Risk of reporting biases in syntheses

The possibility of reporting bias could not be excluded, as not all trials reported all relevant outcomes. The planned funnel plots were not created because we did not include 10 or more studies reporting on any of the outcomes.

### Certainty of evidence

The effect of corticosteroids, compared with placebo or no treatment, is uncertain for maternal death, acute pulmonary edema, dialysis, and perinatal death. We downgraded the certainty of the evidence three levels to very low due to extremely serious imprecision. The 95% CI is very wide and includes both large benefit and large harm. The two boundaries of CI suggest very different inferences.

Corticosteroids, compared with placebo or no treatment, may result in a slight reduction in acute renal failure. We downgraded the certainty of the evidence two levels to low due to very serious imprecision. The 95% CI overlaps no effect and includes large benefit.

Corticosteroids, compared with placebo or no treatment, have little or no difference in the need for platelet transfusion. We downgraded the certainty of evidence two levels to low due to very serious imprecision. The pooled estimate of the risk ratio suggests no difference and the CI includes appreciable benefit and harm.

A Summary of Findings table presents the same information as the text above, with footnotes explaining judgments, Table 2.

## Discussion

We conducted this systematic review and meta-analysis to assess the effects of corticosteroids for improving outcomes in women with HELLP syndrome. This updated evidence synthesis is mandatory for the development of the Egyptian National Guideline for the management of severe preeclampsia, commissioned by the Egyptian Health Council.

### Summary of the evidence

There was no clear evidence of a treatment effect of corticosteroids on substantive clinical outcomes. The effect of corticosteroids, compared with placebo or no treatment, is uncertain for maternal death, acute pulmonary edema, dialysis, and perinatal death. Corticosteroids, compared with placebo or no treatment, have little or no difference in the need for platelet transfusion but may result in a slight reduction in acute renal failure.

The results of this up-to-date review are consistent with the findings reported previously [8] that there was insufficient evidence to support the administration of corticosteroids to women with HELLP syndrome.

In this review, we only included randomized trials for the meta-analysis. A major issue of observational evidence is that it is known to have limited internal validity as it is subject to both bias and confounding, therefore observational studies were excluded to ensure reliability of the results by minimizing the risk of bias due to unmeasured confounders. Overall, observational study designs are not the most appropriate to assess the causal relationship between an intervention and an outcome as several characteristics might differ or might change over time between the different intervention groups. So, the inclusion of observational studies in a meta-analysis might introduce bias in the summary effect. Potential biases are likely to be greater for observational studies compared with randomized trials when evaluating the effects of interventions.

Observational studies of interventions vary in their ability to estimate a causal effect. Biases affecting observational studies of interventions vary depending on the features of the studies. Published reviews [30] that included observational studies have not adequately addressed potential confounders and the likelihood of increased heterogeneity resulting from residual confounding and from other biases that vary across studies.

Our strategy aimed to study the effectiveness of corticosteroids in HELLP syndrome for improving critical maternal and perinatal outcomes rather than surrogate outcomes. While surrogate outcome measures, such as platelet count and liver enzymes laboratory results, may provide insights into how a treatment might work, yet they do not necessarily reflect clinical benefits relevant to decision making. Some interventions that reduce the risk for a surrogate outcome may have no or harmful effects on clinically relevant outcomes, and other interventions having no effect on surrogate measures may improve clinical outcomes. [31,32] As such, and despite their potential appeal, superiority on a surrogate end point, for example the change in platelet count and liver enzymes, may not reflect actual benefits that corticosteroids have on critical outcomes of women with HELLP syndrome. Furthermore, surrogate end points are potentially misleading and should be avoided, or at least interpreted with caution, as decision makers are required to extrapolate the findings to estimate true patient benefits, resulting in uncertainty about the effect of corticosteroids in HELLP syndrome. Published synthesized evidence [30] that included surrogate outcomes without downgrading the certainty of evidence for indirectness would provide misleading implications for practice. In the presence of patient relevant outcomes, the use of surrogate outcomes in a synthesis of evidence to inform practice can not be justified. [33]

We focused on studies that compared corticosteroids to placebo or standard care. Various types of corticosteroids differ in their relative potency and duration of action. It would, therefore, be counter-intuitive, and not clinically useful, to compare one corticosteroid to another when evidence fails to show a difference between any corticosteroid vs placebo or no treatment. Investigators [34] raised serious concerns regarding the credibility of the subgroup analysis results of the Cochrane Review [8] and the application of these subgroup results into clinical practice.

I*n summary*, our methodology minimized bias through strict inclusion of randomized controlled trials, established the class effect first before agent comparisons, and emphasized outcomes of greatest clinical relevance. This approach provided the most robust and applicable evidence for clinical decision making.

The results of our up-to-date synthesis of available evidence provide a rigorous evidence base for trustworthy clinical practice guidelines for the management of HELLP syndrome. [35–41]

### Limitations

A limitation of the evidence was the restricted number of outcomes reported in the included trials. Most included trials reported surrogate laboratory results. The possibility of reporting bias could not be excluded, given that not all trials reported all relevant outcomes. In the case of HELLP syndrome, patient-relevant outcomes do not require exceptional training, expensive tools, or long follow up. It would be implausible to conduct a trial in such a critical condition without reporting maternal death or morbidity.

## Conclusions

In women with HELLP syndrome, the effect of corticosteroids versus placebo or no treatment is uncertain for critical patient-relevant outcomes. The currently available evidence does not support or refute the practice of corticosteroid administration for treating HELLP syndrome.

## Data Availability

All data produced in the present work are contained in the manuscript and its supplemental files.

## Acknowledgments

We acknowledge the contribution and the spirit of goodwill of Dr. Maha Khalifa, the healthcare advocate for her valuable insights.

## Author contributions

AN conceived the idea for this review and designed the review methods. AFK, HBA coordinated the research activities. All authors collaborated in searching, screening, selecting studies, data extraction and synthesis. AFN, AFK, HBA wrote the first draft of the manuscript. All authors revised the manuscript critically for important intellectual content. All authors approved the final version of the manuscript. AFN is the guarantor for this manuscript.

## Availability of data and materials

All data relevant to this study are publicly available. Data, analysis script and materials related to this study are publicly available on the Open Science Framework at https://osf.io/9vwdq. The study protocol and materials were registered on 18 September 2023 at https://osf.io/yzku5. To facilitate reproducibility, this manuscript was written by interleaving regular prose and analysis code using R Markdown.

## Ethics approval and consent to participate

Not applicable.

## Consent for publication

Not applicable.

## Supplemental files

Supplemental file 1

Supplemental file 2

Supplemental file 3

Supplemental file 4

