## Supplemental file 1 for "Corticosteroids for improving patient relevant outcomes in HELLP syndrome: a systematic review and meta-analysis"

#### Abstract

Objective: We will conduct an update of a systematic review and meta-analysis to assess the benefits and harms of corticosteroids vs placebo or no treatment in hemolysis, elevated liver enzymes and low platelets (HELLP) syndrome.

Methods: The search will include bibliographic databases (CENTRAL, MEDLINE, CINAHL), citation indexes (Web of Science and Scopus), and clinical trial registries (ClinicalTrials.gov and the World Health Organization International Clinical Trials Registry Platform). We will search reference lists and explore the cited-by logs of relevant studies and previously published reviews. Randomized trials comparing any corticosteroid with placebo or no treatment in HELLP syndrome will be included. Two authors will assess risk of bias and extract data independently. The main outcome measure is maternal death. We will conduct common-effects meta-analysis where appropriate. We will conduct subgroup analyses by ante- versus postpartum administration and by type of corticosteroid. We will use the Grading of Recommendations, Assessment, Development and Evaluation (GRADE) approach to create the Summary of Findings table.

Discussion: The update is required for the development of the National Guideline for the management of severe preeclampsia. Keywords: Corticosteroids, HELLP Syndrome, Preeclampsia, Maternal morbidity

#### Introduction

The syndrome of hemolysis, elevated liver enzymes and low platelets (HELLP) has an incidence of 2.5 per 1000 singleton deliveries and It complicates 20% of severe pre-eclampsia. (1,2)

The pathophysiology of HELLP syndrome that is usually diagnosed between 27 and 37 weeks, is not completely understood. (3)

The diagnosis of HELLP syndrome depends on laboratory findings of microangiopathic hemolysis, thrombocytopenia, and elevated liver enzymes. Different investigators reported different threshold of hematologic and biochemical values for diagnosis of the syndrome or for determining the prognosis. (4–6)

The presence of HELLP syndrome is associated with significant maternal mortality and morbidity including acute renal and liver failure. (1)

Approximately 70% of pregnancies complicated by HELLP syndrome require preterm delivery, thus increasing perinatal morbidity and mortality. (7)

Observational studies suggested that steroid treatment in HELLP syndrome may improve disordered maternal hematological and biochemical features and perhaps perinatal mortality and morbidity. Clinical trials examined the effects of corticosteroids for the treatment of maternal HELLP syndrome. Various regimens have been reported using prednisolone, dexamethasone, or betamethasone. (2,8)

Current practice and clinical guidelines require an updated evidence synthesis as the available Cochrane review was published in 2010. (9)

We will conduct this updated systematic review and meta-analysis to assess the benefits and harms of corticosteroids versus placebo or no treatment in women with HELLP syndrome.

### Methods

Protocol and registration

We prepared the protocol following the methodological standards of Cochrane handbook. (10) We prospectively registered the protocol in Open Science Platform. The full text of the protocol is available in an open access registry.

We reported the protocol using the Preferred Reporting Items for Systematic reviews and Meta-Analyses-Protocol (PRISMA-P) standards. (11)

Eligibility criteria

We will include published randomized controlled trials that recruited women with HELLP syndrome, confirmed by objective testing. We will include studies comparing corticosteroids versus placebo or no intervention. The outcome measures include maternal death, severe maternal morbidity, admission to intensive care unit, use of mechanical ventilation, dialysis, postpartum hemorrhage defined as blood loss of 1000 mL or greater, liver hematoma, ruptured liver, need for platelet transfusion, perinatal death, and severe perinatal morbidity.

Information sources

A comprehensive literature search will be initially conducted on August 31, 2023. We will not impose language or other restrictions on any of the searches. We will search bibliographic databases (Cochrane Central Register of Controlled Trials (CENTRAL), MEDLINE, CINAHL), and citation indexes (Web of Science and Scopus). We will search clinical trial registries (ClinicalTrials.gov and the World Health Organization International Clinical Trials Registry Platform) to identify ongoing trials. We will search reference lists and explore the cited-by logs of identified studies and previously published reviews.

The search strategy is designed by a search expert with input from the authors. We will use the following search strategy for MEDLINE/PubMed.

#1 "randomized controlled trial"[pt]
#2 "controlled clinical trial"[pt]
#3 randomized[tiab]
#4 placebo[tiab]
#5 "clinical trials as topic"[Mesh:NoExp]
#6 randomly[tiab]
#7 trial[ti]
#8 #1 OR #2 OR #3 OR #4 OR #5 OR #6 OR #7
#9 animals[Mesh] NOT humans[Mesh]

| | |
|---|---|
| #10 | #8 NOT #9 |
| #11 | "HELLP Syndrome"[Mesh] |
| #12 | "HELLP Syndrome"[tiab] |
| #13 | #11 OR #12 |
| #14 | Steroids[Mesh] |
| #15 | Glucocorticoids[PA] |
| #16 | Dexamethasone[tiab] |
| #17 | Betamethasone[tiab] |
| #18 | Prednisolone[tiab] |
| #19 | #14 OR #15 OR #16 OR #17 OR #18 |
| #20 | #10 AND #13 AND #19 |

The detailed exact strategy adapted for each database is provided in Appendix 1 and an open-access registry document. (12)

Study selection

Two authors will independently screen all titles and abstracts for eligibility. We will retrieve and assess the full text of all the studies that potentially meet our eligibility criteria during screening. Two authors will independently assess each full-text article based on the eligibility criteria described above. Disagreements regarding trial eligibility will be resolved by consensus and finally resolved by a third author (AN).

Data collection process

For eligible studies, two authors will extract the data in duplicates using an offline electronic form. We will resolve discrepancies through discussion. We will enter the data into a spreadsheet and check them for accuracy. We will contact authors of the original reports to provide further details regarding unclear or missing data.

Data items

We will extract study design, description of included participants, description of the intervention and comparators, outcomes, trial registration, and funding sources.

Risk of bias in individual studies

We will assess the risk of bias using Cochrane RoB-2 (Appendix 2). We will use the criteria recently outlined in the Cochrane Handbook for Systematic Reviews of Interventions. (10) Two authors will independently assess the risk of bias in each trial. We will resolve any differences of opinion regarding assessment of risk of bias by discussion.

Summary measures

An intent-to-treat analysis, including all randomized women, will be performed. We do not expect to include any cluster, or crossover trials. For dichotomous data, we will present results as summary risk ratio (RR) with 95% confidence intervals (CI). For continuous data we will present results as summary mean difference (MD) with 95% CI.

Synthesis of results

Fixed-effect meta-analysis will be performed to combine data of trials that are judged to be sufficiently similar in terms of intervention, populations, and methods. Substantial statistical heterogeneity, defined as $I^2$ statistic ≥ 50% or $P < 0.1$, will be investigated and a random-effects meta-analysis will be performed only if an average treatment effect across trials was considered clinically meaningful. The number needed to treat (NNT) for benefit or harm with the 95% CI will be calculated for outcomes for which there is a statistically significant difference.

Meta-analysis will be performed using R software version 4.3 (13) and package meta version 6.5. (14)

Meta-bias

The assessment of publication bias will be assessed using a funnel plot and standard tests.

Additional analyses

Substantial heterogeneity will be thoroughly investigated based on the prespecified methods. We will perform the planned subgroup analysis by gestational age at enrollment (ante- vs postpartum) and by class of corticosteroids. We will assess subgroup differences by interaction tests available within R. Results of the subgroup analyses were reported by mentioning the $Chi^2$ statistic and P value, and the interaction test $I^2$ value.

Sensitivity analyses will be performed to explore robustness of pooled estimate using random effect model and fixed effect model for the outcome of continuation. We will also conduct sensitivity analyses to explore the effects of incomplete outcome data by conducting an available case versus a worst-case scenario analysis to evaluate robustness of results.

Summary of findings

We will use the Grading of Recommendations, Assessment, Development and Evaluation (GRADE) approach to create the Summary of findings table. (15) Briefly, GRADE uses study limitations, consistency of effect, imprecision, indirectness, and publication bias to assess the quality of the body of evidence for each outcome. A summary of the intervention effect and a measure of quality for outcomes will be produced using the GRADEpro GDT software. (16) One author (A.N.) will conduct GRADE assessments and the decisions on downgrading. This will be discussed for final approval by all authors.

### Discussion

Current practice and clinical guidelines require an updated evidence synthesis as the available Cochrane review was published in 2010 and several trials have been published later. We will conduct this updated systematic review and meta-analysis to assess the benefits and harms of corticosteroids for improving maternal outcomes in women with HELLP syndrome. The update is required for the development of the National Guideline for the management of severe preeclampsia.

### Appendix 1: Search strategies

Date: Wednesday, August 30, 2023

#### PubMed

"randomized controlled trial"[Publication Type]
"controlled clinical trial"[Publication Type]
randomized[Title/Abstract]
placebo[Title/Abstract]
"clinical trials as topic"[MeSH Terms:no exp]
randomly[Title/Abstract]
trial[Title]
#1 OR #2 OR #3 OR #4 OR #5 OR #6 OR #7
animals[MeSH Terms] NOT humans[MeSH Terms]
#8 NOT #9
"HELLP Syndrome"[MeSH Terms]
"HELLP Syndrome"[Title/Abstract]
#11 OR #12
Steroids[MeSH Terms]
Glucocorticoids[Pharmacological Action]
Dexamethasone[Title/Abstract]
Betamethasone[Title/Abstract]
Prednisolone[Title/Abstract]
#14 OR #15 OR #16 OR #17 OR #18
#10 AND #13 AND #19

#### Web of Science

DT="randomized controlled trial"
DT="controlled clinical trial"
(TI=randomized OR AB=randomized)
(TI=placebo OR AB=placebo)
MH="clinical trials as topic"
(TI=randomly OR AB=randomly)
TI=trial
#1 OR #2 OR #3 OR #4 OR #5 OR #6 OR #7
MHX=animals NOT MHX=humans
#8 NOT #9
MHX="HELLP Syndrome"
(TI="HELLP Syndrome" OR AB="HELLP Syndrome")
#11 OR #12
MHX=Steroids
MHX=Glucocorticoids
(TI=Dexamethasone OR AB=Dexamethasone)
(TI=Betamethasone OR AB=Betamethasone)
(TI=Prednisolone OR AB=Prednisolone)
#14 OR #15 OR #16 OR #17 OR #18
#10 AND #13 AND #19

#### CENTRAL

#1 ("HELLP syndrome"):ti,ab,kw
#2 MeSH descriptor: [HELLP Syndrome] explode all trees
#3 #1 OR #2
#4 MeSH descriptor: [Steroids] explode all trees
#8 MeSH descriptor: [Glucocorticoids] explode all trees
#5 (Dexamethasone):ti,ab,kw
#6 (Betamethasone):ti,ab,kw
#7 ("prednisolone"):ti,ab,kw
#9 #4 OR #5 OR #6 OR #7 OR #8
#10 #3 AND #9 in Trials

#### Scopus

INDEXTERMS ("randomized controlled trial")
INDEXTERMS ("controlled clinical trial")
INDEXTERMS("clinical trials as topic")

TITLE-ABS(randomized)
TITLE-ABS(placebo)
TITLE-ABS(randomly)
TITLE(trial)
#1 OR #2 OR #3 OR #4 OR #5 OR #6 OR #7
INDEXTERMS(animals) AND NOT INDEXTERMS(humans)
#8 AND NOT #9
INDEXTERMS("HELLP Syndrome")
TITLE-ABS("HELLP Syndrome")
#11 OR #12
INDEXTERMS(Steroids)
INDEXTERMS(Glucocorticoids)
TITLE-ABS(Dexamethasone)
TITLE-ABS(Betamethasone)
TITLE-ABS(Prednisolone)
#14 OR #15 OR #16 OR #17 OR #18
#10 AND #13 AND #19

#### **Appendix 1:** Risk-of-bias 2 tool (RoB 2)

**Domain 1: Risk of bias arising from the randomization process.**

| Signaling questions | Comments | Response options |
|---|---|---|
| 1.1 Was the allocation sequence random? | | Y / PY / PN / N / NI |
| 1.2 Was the allocation sequence concealed until participants were enrolled and assigned to interventions? | | Y / PY / PN / N / NI |
| 1.3 Did baseline differences between intervention groups suggest a problem with the randomization process? | | Y / PY / PN / N / NI |
| Risk-of-bias judgement | | Low / High / Some concerns |

**Domain 2: Risk of bias due to deviations from the intended interventions.**

| Signalling questions | Comments | Response options |
|---|---|---|
| 2.1. Were participants aware of their assigned intervention during the trial? | | Y / PY / PN / N / NI |
| 2.2. Were carers and people delivering the interventions aware of participants' assigned intervention during the trial? | | Y / PY / PN / N / NI |
| 2.3. If Y/PY/NI to 2.1 or 2.2: Were there deviations from the intended intervention that arose because of the trial context? | | NA / Y / PY / PN / N / NI |
| 2.4 If Y/PY to 2.3: Were these deviations likely to have affected the outcome? | | NA / Y / PY / PN / N / NI |
| 2.5. If Y/PY/NI to 2.4: Were these deviations from intended intervention balanced between groups? | | NA / Y / PY / PN / N / NI |
| 2.6 Was an appropriate analysis used to estimate the effect of intervention? | | Y / PY / PN / N / NI |
| 2.7 If N/PN/NI to 2.6: Was there potential for a substantial impact (on the result) of the failure to analyse participants in the group to which they were randomized? | | NA / Y / PY / PN / N / NI |
| Risk-of-bias judgement | | Low / High / Some concerns |

**Domain 3: Missing outcome data**

| Signaling questions | Comments | Response options |
|---|---|---|
| 3.1 Were data for this outcome available for all, or nearly all, participants randomized? | | Y / PY / PN / N / NI |
| 3.2 If N/PN/NI to 3.1: Is there evidence that the result was not biased by missing outcome data? | | NA / Y / PY / PN / N |
| 3.3 If N/PN to 3.2: Could missingness in the outcome depend on its true value? | | NA / Y / PY / PN / N/NI |
| 3.4 If Y/PY/NI to 3.3: Is it likely that missingness in the outcome depended on its true value? | | NA / Y / PY / PN / N/NI |
| Risk-of-bias judgement | | Low / High / Some concerns |

##### Domain 4: Risk of bias in measurement of the outcome

| Signalling questions | Comments | Response options |
|---|---|---|
| 4.1 Was the method of measuring the outcome inappropriate? | | Y / PY / PN / N / NI |
| 4.2 Could measurement or ascertainment of the outcome have differed between intervention groups? | | Y / PY / PN / N / NI |
| 4.3 If N/PN/NI to 4.1 and 4.2: Were outcome assessors aware of the intervention received by study participants? | | NA / Y / PY / PN / N / NI |
| 4.4 If Y/PY/NI to 4.3: Could assessment of the outcome have been influenced by knowledge of intervention received? | | NA / Y / PY / PN / N / NI |
| 4.5 If Y/PY/NI to 4.4: Is it likely that assessment of the outcome was influenced by knowledge of intervention received? | | NA / Y / PY / PN / N / NI |
| Risk-of-bias judgement | | Low / High / Some concerns |

##### Domain 5: Risk of bias in selection of the reported result

| Signalling questions | Comments | Response options |
|---|---|---|
| 5.1 Were the data that produced this result analysed in accordance with a pre-specified analysis plan that was finalized before unblinded outcome data were available for analysis? | | Y / PY / PN / N / NI |
| Is the numerical result being assessed likely to have been selected, on the basis of the results, from... | | |
| 5.2. ... multiple eligible outcome measurements (e.g. scales, definitions, time points) within the outcome domain? | | Y / PY / PN / N / NI |
| 5.3 ... multiple eligible analyses of the data? | | Y / PY / PN / N / NI |
| Risk-of-bias judgement | | Low / High / Some concerns |

#### Overall risk of bias

| | | |
|---|---|---|
| Overall risk-of-bias judgement | | Low / High / Some concerns |
