## Supplemental file 4 for "Corticosteroids for improving patient relevant outcomes in HELLP syndrome: a systematic review and meta-analysis"

### Supplementary file 4 Data and synthesis script

2023-09-26

Dataset

| id | ee | ne | ec | nc | factor | corticosteroid | outcome | RoB | zero |
|---|---|---|---|---|---|---|---|---|---|
| Van Runnard 2006 | 1 | 15 | 1 | 16 | antepartum | prednisolone | Abruptio placenta | low | F |
| Magann 1994 | 3 | 12 | 3 | 13 | antepartum | dexa | Apgar score at 5 minutes < 7 | some concerns | F |
| Van Runnard 2006 | 1 | 16 | 2 | 17 | antepartum | prednisolone | Apgar score at 5 minutes < 7 | low | F |
| Ozer 2009 | 23 | 30 | 26 | 30 | antepartum | beta | Cesarean section | low | F |
| Van Runnard 2006 | 15 | 15 | 14 | 16 | antepartum | prednisolone | Cesarean section | low | F |
| Fonseca 2019 | 6 | 35 | 8 | 38 | mixed | dexa | Composite morbidity | low | F |
| Van Runnard 2006 | 1 | 15 | 4 | 16 | antepartum | prednisolone | Composite morbidity | low | F |
| Ozer 2009 | 1 | 30 | 0 | 30 | antepartum | beta | Dialysis | low | F |
| Fonseca 2005 | 8 | 66 | 10 | 66 | mixed | dexa | Eclampsia | low | F |
| Fonseca 2019 | 2 | 40 | 1 | 41 | mixed | dexa | Eclampsia | low | F |
| Magann 1994 | 1 | 12 | 0 | 13 | antepartum | dexa | ICH | some concerns | F |
| Van Runnard 2006 | 4 | 16 | 2 | 17 | antepartum | prednisolone | ICH | low | F |
| Fonseca 2005 | 3 | 66 | 1 | 66 | mixed | dexa | Maternal death | low | F |
| Fonseca 2019 | 0 | 42 | 2 | 45 | mixed | dexa | Maternal death | low | F |
| Katz 2008 | 2 | 56 | 2 | 49 | postpartum | dexa | Maternal death | low | F |
| Ozer 2009 | 0 | 30 | 0 | 30 | antepartum | beta | Maternal death | low | T |
| Van Runnard 2006 | 0 | 15 | 1 | 16 | antepartum | prednisolone | Maternal death | low | F |
| Vigil-De Gracia 1997 | 0 | 17 | 1 | 17 | postpartum | dexa | Maternal death | high | F |
| Ozer 2009 | 0 | 30 | 1 | 30 | antepartum | beta | Maternal liver morbidity | low | F |
| Van Runnard 2006 | 0 | 15 | 3 | 16 | antepartum | prednisolone | Maternal liver morbidity | low | F |
| Fonseca 2005 | 3 | 66 | 1 | 66 | mixed | dexa | Maternal pulmonary edema | low | F |
| Fonseca 2019 | 0 | 41 | 1 | 43 | mixed | dexa | Maternal pulmonary edema | low | F |
| Katz 2008 | 2 | 56 | 5 | 49 | postpartum | dexa | Maternal pulmonary edema | low | F |

| | | | | | | | | | |
|---|---|---|---|---|---|---|---|---|---|
| Ozer 2009 | 1 | 30 | 1 | 30 | antepartum | beta | Maternal pulmonary edema | low | F |
| Fonseca 2005 | 6 | 66 | 8 | 66 | mixed | dexa | Maternal renal failure | low | F |
| Fonseca 2019 | 1 | 37 | 6 | 42 | mixed | dexa | Maternal renal failure | low | F |
| Katz 2008 | 9 | 56 | 12 | 49 | postpartum | dexa | Maternal renal failure | low | F |
| Ozer 2009 | 2 | 30 | 3 | 30 | antepartum | beta | Maternal renal failure | low | F |
| Yalcin 1998 | 3 | 15 | 3 | 15 | postpartum | dexa | Maternal renal failure | some concerns | F |
| Van Runnard 2006 | 0 | 16 | 2 | 17 | antepartum | prednisolone | Necrotizing enterocolitis | low | F |
| Magann 1994 | 3 | 12 | 1 | 13 | antepartum | dexa | Neonatal RDS | some concerns | F |
| Van Runnard 2006 | 6 | 16 | 8 | 17 | antepartum | prednisolone | Neonatal RDS | low | F |
| Magann 1994 | 1 | 12 | 3 | 13 | antepartum | dexa | Perinatal death | some concerns | F |
| Van Runnard 2006 | 3 | 16 | 4 | 17 | antepartum | prednisolone | Perinatal death | low | F |
| Fonseca 2005 | 12 | 66 | 10 | 66 | mixed | dexa | Platelet transfusion | low | F |
| Fonseca 2019 | 12 | 42 | 15 | 45 | mixed | dexa | Platelet transfusion | low | F |

#### Synthesis

##### Pre-specified outcomes

1. Maternal Death
2. Liver morbidity (hematoma, rupture, failure)
3. Acute pulmonary edema
4. Acute renal failure
5. Dialysis
6. Platelet transfusion
7. Perinatal death

Maternal Death

Pairwise meta-analysis

| Study | RR | 95%-CI % | W(common) |
|---|---|---|---|
| Fonseca 2005 | 3.0000 | [0.3202; 28.1042] | 25.4 |
| Fonseca 2019 | 0.2141 | [0.0106; 4.3328] | 14.1 |
| Katz 2008 | 0.8750 | [0.1280; 5.9809] | 34.5 |
| Ozer 2009 | NA | | 0.0 |
| Van Runnard 2006 | 0.3548 | [0.0156; 8.0730] | 13.0 |
| Vigil-De Gracia 1997 | 0.3333 | [0.0146; 7.6344] | 13.0 |

Number of studies: k = 5

Number of observations: o = 449

Number of events: e = 12

| | RR | 95%-CI | z | p-value |
|---|---|---|---|---|
| Common effect model | 0.7700 | [0.2492; 2.3799] | -0.45 | 0.6499 |

Quantifying heterogeneity:

tau^2 = 0 [0.0000; 7.4236]; tau = 0 [0.0000; 2.7246]

I^2 = 0.0% [0.0%; 79.2%]; H = 1.00 [1.00; 2.19]

Test of heterogeneity: Q = 2.64; d.f. = 4; p-value 0.6192

Details on meta-analytical method:

- Inverse variance method
- Restricted maximum-likelihood estimator for tau^2
- Q-Profile method for confidence interval of tau^2 and tau
- Continuity correction of 0.5 in studies with zero cell frequencies

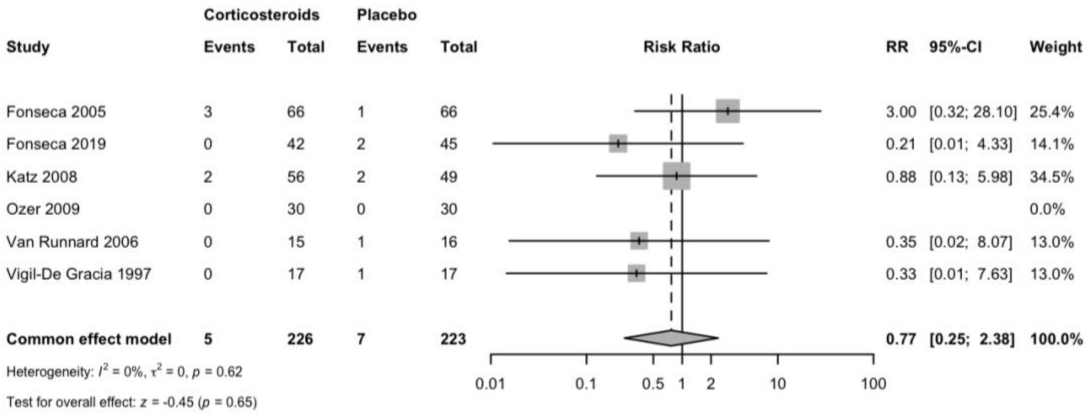

#### Risk of bias

##### Summary traffic-light plot of risk-of-bias assessments

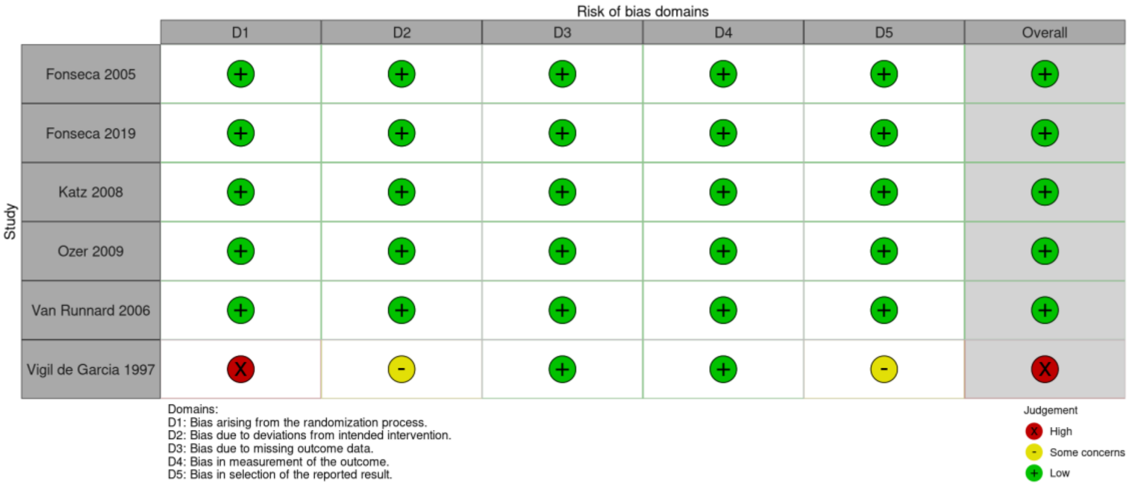


##### Summary weighted barplot of risk-of-bias assessments

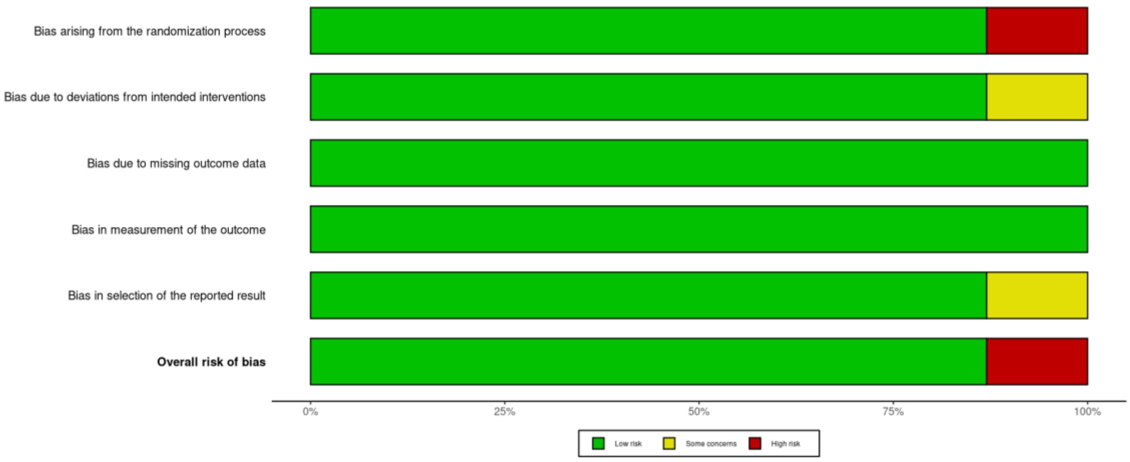

Subgroup analysis: Antepartum vs postpartum vs mixed

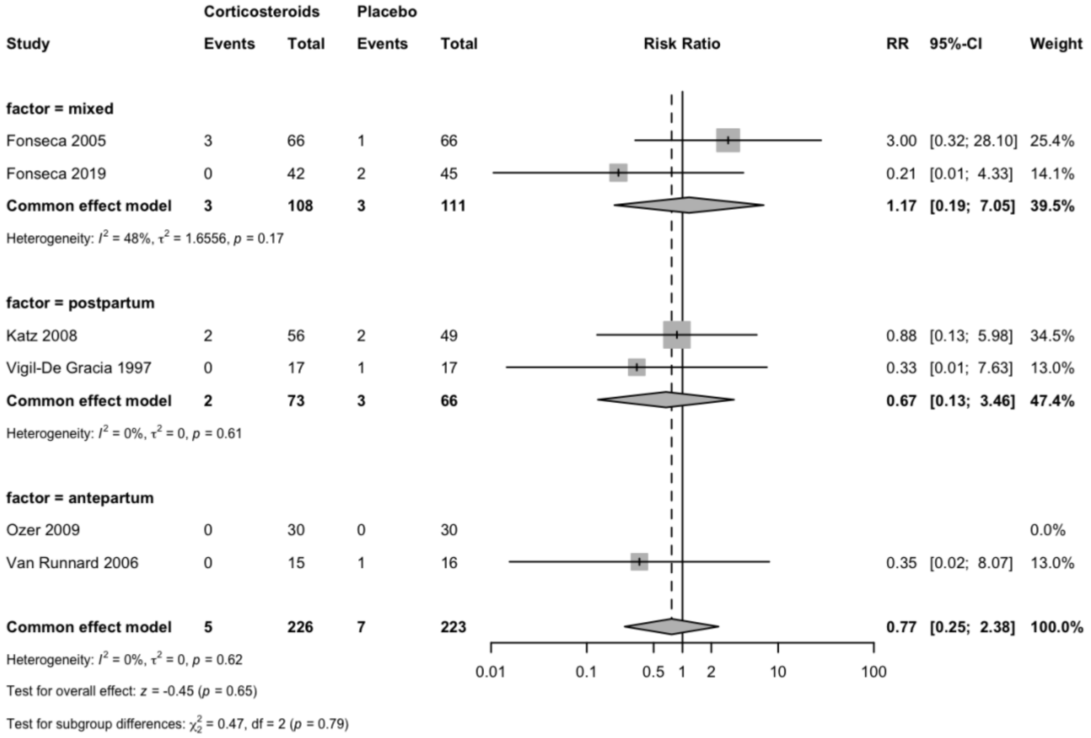


Subgroup analysis: Dexa vs Beta vs Prednisolone

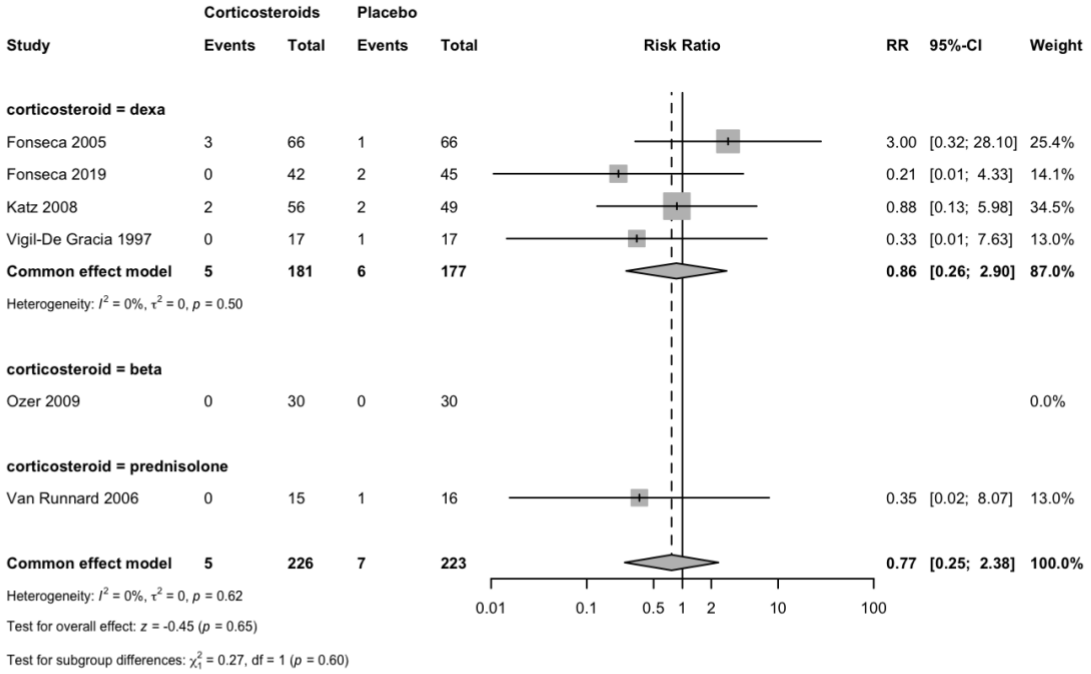

Sensitivity analysis: Low RoB

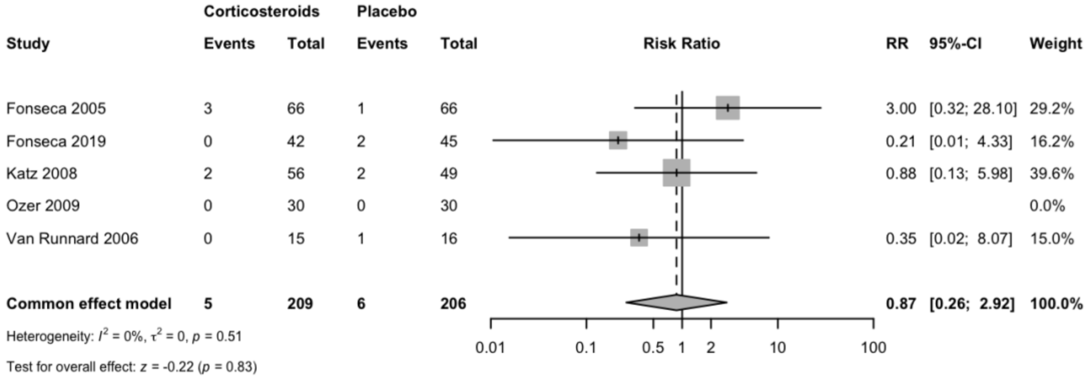


Sensitivity analysis: Studies with zero events included

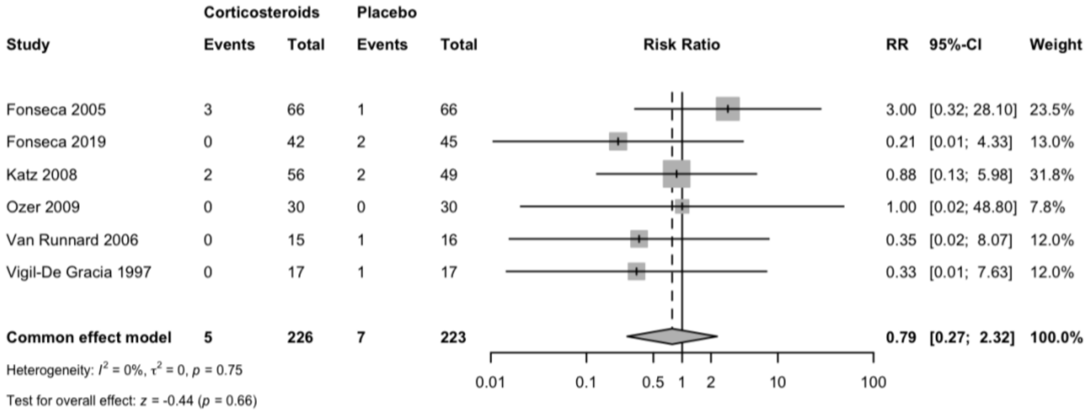


Funnel plot

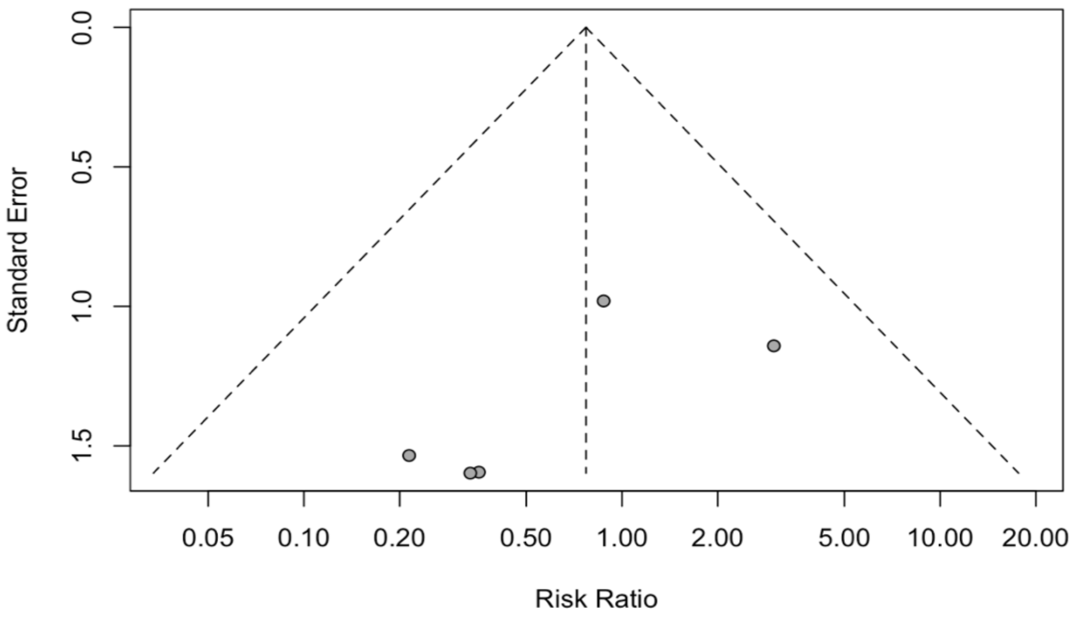

Acute pulmonary edema

RR 95%-CI %W(common)

Fonseca 2005 3.0000 [0.3202; 28.1042] 24.2

Fonseca 2019 0.3494 [0.0146; 8.3368] 12.0

Katz 2008 0.3500 [0.0711; 1.7238] 47.6

Ozer 2009 1.0000 [0.0655; 15.2598] 16.3

Number of studies: k = 4

Number of observations: o = 381

Number of events: e = 14

RR 95%-CI z p-value

Common effect model 0.6975 [0.2323; 2.0944] -0.64 0.5207

Quantifying heterogeneity:

tau^2 = 0.1164 [0.0000; 13.2337]; tau = 0.3412 [0.0000; 3.6378]

I^2 = 0.0% [0.0%; 84.7%]; H = 1.00 [1.00; 2.56]

Test of heterogeneity:

Q d.f. p-value

2.60 3 0.4572

Details on meta-analytical method:

- Inverse variance method
- Restricted maximum-likelihood estimator for tau^2
- Q-Profile method for confidence interval of tau^2 and tau
- Continuity correction of 0.5 in studies with zero cell frequencies

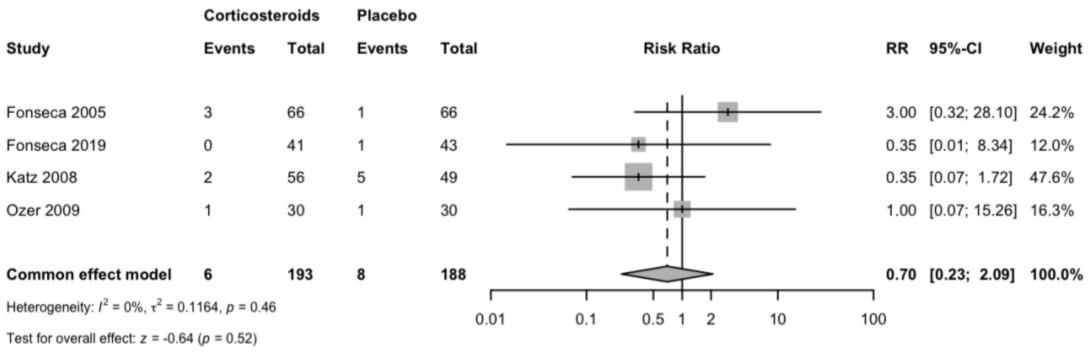

Acute renal failure

| Study | RR | 95%-CI % | W(common) |
|---|---|---|---|
| Fonseca 2005 | 0.7500 | [0.2754; 2.0428] | 26.7 |
| Fonseca 2019 | 0.1892 | [0.0239; 1.4998] | 6.3 |
| Katz 2008 | 0.6563 | [0.3025; 1.4238] | 44.8 |
| Ozer 2009 | 0.6667 | [0.1198; 3.7087] | 9.1 |
| Yalcin 1998 | 1.0000 | [0.2390; 4.1844] | 13.1 |

Number of studies: k = 5
Number of observations: o = 406
Number of events: e = 53

| | RR | 95%-CI | z | p-value |
|---|---|---|---|---|
| Common effect model | 0.6658 | [0.3965; 1.1179] | -1.54 | 0.1239 |

Quantifying heterogeneity:

tau^2 = 0 [0.0000; 2.4607]; tau = 0 [0.0000; 1.5687]
I^2 = 0.0% [0.0%; 79.2%]; H = 1.00 [1.00; 2.19]
Test of heterogeneity: Q = 1.78 ; d.f. = 4; p-value = 0.7753

Details on meta-analytical method:

- Inverse variance method
- Restricted maximum-likelihood estimator for tau^2
- Q-Profile method for confidence interval of tau^2 and tau

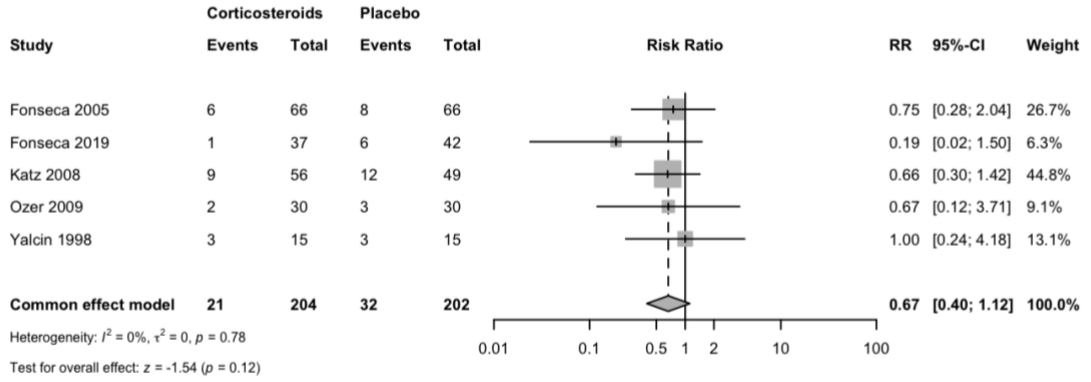


Dialysis

Number of observations: o = 60
Number of events: e = 1

| Study | RR | 95%-CI | z | p-value |
|---|---|---|---|---|
| Ozer 2009 | 3.0000 | [0.1271; 70.7833] | 0.68 | 0.4958 |

Details: Continuity correction of 0.5

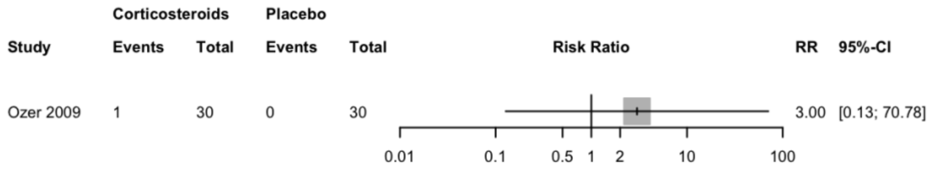

Liver morbidity

```
                 RR          95%-CI %W(common)
Ozer 2009        0.3333 [0.0141; 7.8648]       45.4
Van Runnard 2006 0.1521 [0.0085; 2.7116]       54.6

Number of studies: k = 2
Number of observations: o = 91
Number of events: e = 4

                    RR          95%-CI     z p-value
Common effect model 0.2171 [0.0258; 1.8257] -1.41  0.1598

Quantifying heterogeneity:
 tau^2 = 0; tau = 0; I^2 = 0.0%; H = 1.00

Test of heterogeneity:
    Q d.f. p-value
 0.13    1  0.7191

Details on meta-analytical method:
- Inverse variance method
- Restricted maximum-likelihood estimator for tau^2
- Continuity correction of 0.5 in studies with zero cell frequencies
```

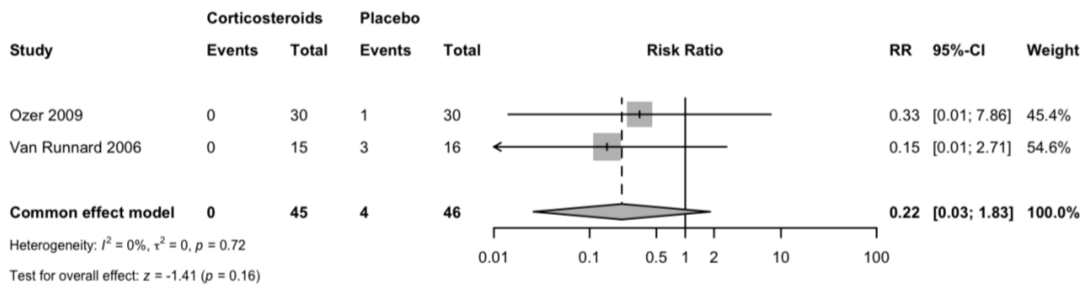

Platelet transfusion

```
              RR        95%-CI %W(common)
Fonseca 2005 1.2000 [0.5574; 2.5832]      40.5
Fonseca 2019 0.8571 [0.4556; 1.6126]      59.5

Number of studies: k = 2
Number of observations: o = 219
Number of events: e = 49

                    RR         95%-CI     z p-value
Common effect model 0.9821 [0.6031; 1.5994] -0.07  0.9422

Quantifying heterogeneity:
 tau^2 = 0; tau = 0; I^2 = 0.0%; H = 1.00

Test of heterogeneity:
    Q d.f. p-value
 0.44    1  0.5069

Details on meta-analytical method:
- Inverse variance method
- Restricted maximum-likelihood estimator for tau^2
```

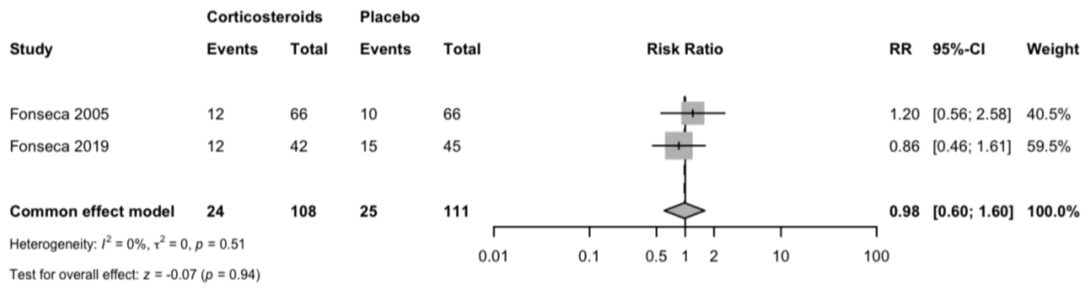

Perinatal death

```
                 RR          95%-CI %W(common)
Magann 1994      0.3611 [0.0432; 3.0169]       28.3
Van Runnard 2006 0.7969 [0.2103; 3.0197]       71.7

Number of studies: k = 2
Number of observations: o = 58
Number of events: e = 11

                    RR           95%-CI     z p-value
Common effect model 0.6372 [0.2062; 1.9693] -0.78  0.4337

Quantifying heterogeneity:
 tau^2 = 0; tau = 0; I^2 = 0.0%; H = 1.00

Test of heterogeneity:
    Q d.f. p-value
 0.38    1  0.5359

Details on meta-analytical method:
- Inverse variance method
- Restricted maximum-likelihood estimator for tau^2
```

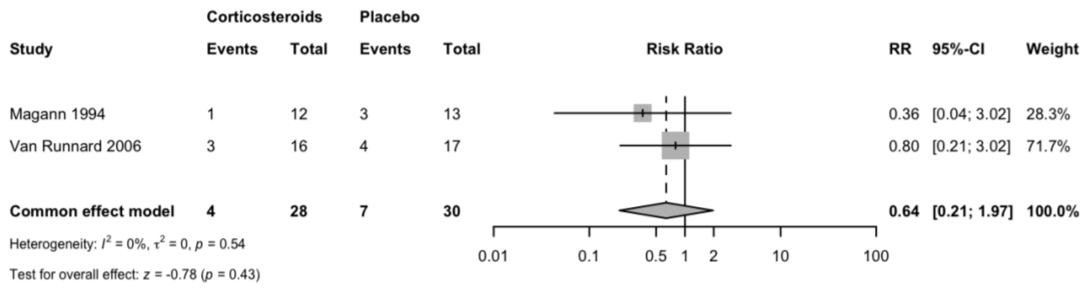
